# Predicting visual function before glaucoma onset from baseline optical coherence tomography scans using deep learning

**DOI:** 10.64898/2026.02.27.26347297

**Authors:** Abadh K Chaurasia, Chen Wang, Patrick W Toohey, Christine Y Chen, Stuart MacGregor, Matthew T Bennett, Nitin Verma, Jamie E Craig, Paul J McCartney, Marc G Sarossy, Alex W Hewitt

## Abstract

**Background:** The visual field (VF) test results of many eyes with glaucoma progress despite treatment. This suggests that some eyes are either untreated or that the management of intraocular pressure (IOP) does not influence the outcome. In this work, we explore whether future VF parameters can be predicted from a baseline optical coherence retinal nerve fibre layer (OCT-RNFL) scan using a deep learning model.

**Methods:** The model was developed using 1792 eyes from 1610 patients, and externally validated on 151 eyes from a second centre using the same Zeiss Cirrus machine and 281 eyes from a third centre using scans obtained from a different (Heidelberg Spectralis) machine. The Vision Transformers (ViT)-based regression model was trained on baseline OCT-RNFL scans to predict three key VF indices (follow-up interval: 4.74 ± 2.59 years). Model performance was evaluated using Mean Absolute Error (MAE) and Root Mean Square Error (RMSE), with 95% confidence intervals (CI).

**Results:** The model achieved an overall MAE of 2.07 (95% CI: 1.91-2.22) and RMSE of 2.87 (95% CI: 2.60-3.14) on the internal validation set. On external validation, the model showed comparable performance with an MAE of 2.07 (95% CI: 1.8-2.35) for the external validation (Zeiss OCT) cohort and 2.11 (95% CI: 1.93-2.31) for the external validation (Heidelberg OCT) cohort. Saliency maps revealed that the inner and outer RNFL layers were key structures in driving the model’s predictions.

**Conclusions:** Our ViT-based regression model effectively predicts key VF indices objectively from a single OCT-RNFL scan, with strong performance across two OCT devices, offering a novel tool for predicting glaucoma progression.

## INTRODUCTION

Glaucoma is a complex chronic optic neuropathy characterised by the degeneration of retinal ganglion cells and their axons, leading to irreversible visual field (VF) loss and long-term functional disability if left untreated.^1,2^ Although intraocular pressure (IOP) lowering remains the only established adjustable risk factor, a substantial number of eyes continue to demonstrate VF progression despite treatment that appears clinically appropriate.^2–4^

Objective structural imaging plays an important role in evaluating glaucomatous damage, especially by assessing the retinal nerve fibre layer (RNFL) with optical coherence tomography (OCT).^1,2^ OCT-derived measures (e.g., peripapillary RNFL) capture glaucomatous neuroretinal change and are routinely acquired in clinics; however, translating baseline structural information into an individual patient’s future functional vision trajectory remains challenging using conventional modelling. Prior machine-learning and deep-learning studies have primarily focused on cross-sectional structure–function mapping, demonstrating that OCT can reconstruct VF patterns or estimate global VF indices: Mean Deviation (MD), Pattern Standard Deviation (PSD), and Visual Field Index (VFI), highlighting the potential to learn structure-function relationships directly from imaging data.^3–5^ Deep learning models can now predict visual field progression using baseline and/or longitudinal structural measurements, such as a series of optic disc photographs and OCT scans.^6–8^ However, while prior studies have estimated global visual field indices from OCT-RNFL and predicted progression risk, direct forecasting of future MD, PSD, and VFI from a single baseline OCT scan remains underexplored.

Although convolutional neural network (CNNs)-based models have traditionally been the dominant deep learning architecture for diagnosing glaucoma using imaging modalities, recent studies suggest that transformer-based models can achieve or exceed the performance of traditional CNNs.^9–11^ CNNs process images through layers that use small spatial filters to detect local patterns, such as edges and textures.^11^ These layers refine features into more abstract representations. Afterwards, two-dimensional (2D) feature maps are flattened into a 1D vector, which passes through fully connected layers to produce a numeric prediction (regressor) or a probability-based decision (classifier). In contrast, a Vision Transformer (ViT) replaces convolutional feature extraction with a patch-based representation and self-attention: the OCT image is divided into fixed-size patches that become a sequence of embedded tokens, and attention learns how to integrate information across the entire image, explicitly modelling long-range spatial relationships rather than relying primarily on local receptive fields.^12,13^ The resulting global embedding is then input to a regression head to produce predictions of MD, PSD, and VFI, providing a transparent 2D-to-1D-to-scalar pathway aligned with our aim of forecasting clinically interpretable global VF indices. In the existing literature, several studies have used CNNs-based regression analyses to estimate VF indices from OCT scans of the ONH, but no previous study has reported forecasting global VF indices before disease worsening using a single baseline OCT-RNFL circular B-scan using a ViT-based regression model.^14–17^

## METHODS

### Study Design

This retrospective, multicentre cohort study used data collected between 2009 and 2025 from three Australian ophthalmology clinics: Hobart Eye Surgeons, Essendon Eye Clinic, and Gladstone Park Eye Clinic. The primary model was trained and internally validated using the Hobart Eye Surgeons dataset (hereafter referred to as the model development cohort), acquired on Carl Zeiss OCT devices. External validation was performed using two independent datasets from Essendon Eye Clinic (Zeiss OCT) and Gladstone Park Eye Clinic (Heidelberg OCT). Data collection at Hobart Eye Surgeons was approved by the University of Tasmania Human Research Ethics Committee (HREC) (29775), and data collection at Essendon Eye Clinic and Gladstone Park Eye Clinic was approved by the Royal Victorian Eye and Ear Hospital HREC (24/1597HL).

Consent waivers were obtained for each of the three clinical sites. In this study, we only extracted patients’ data listed as having glaucoma or being a suspect in the clinic database. An overview of this study is outlined in **Figure 1**.

**Figure 1:**
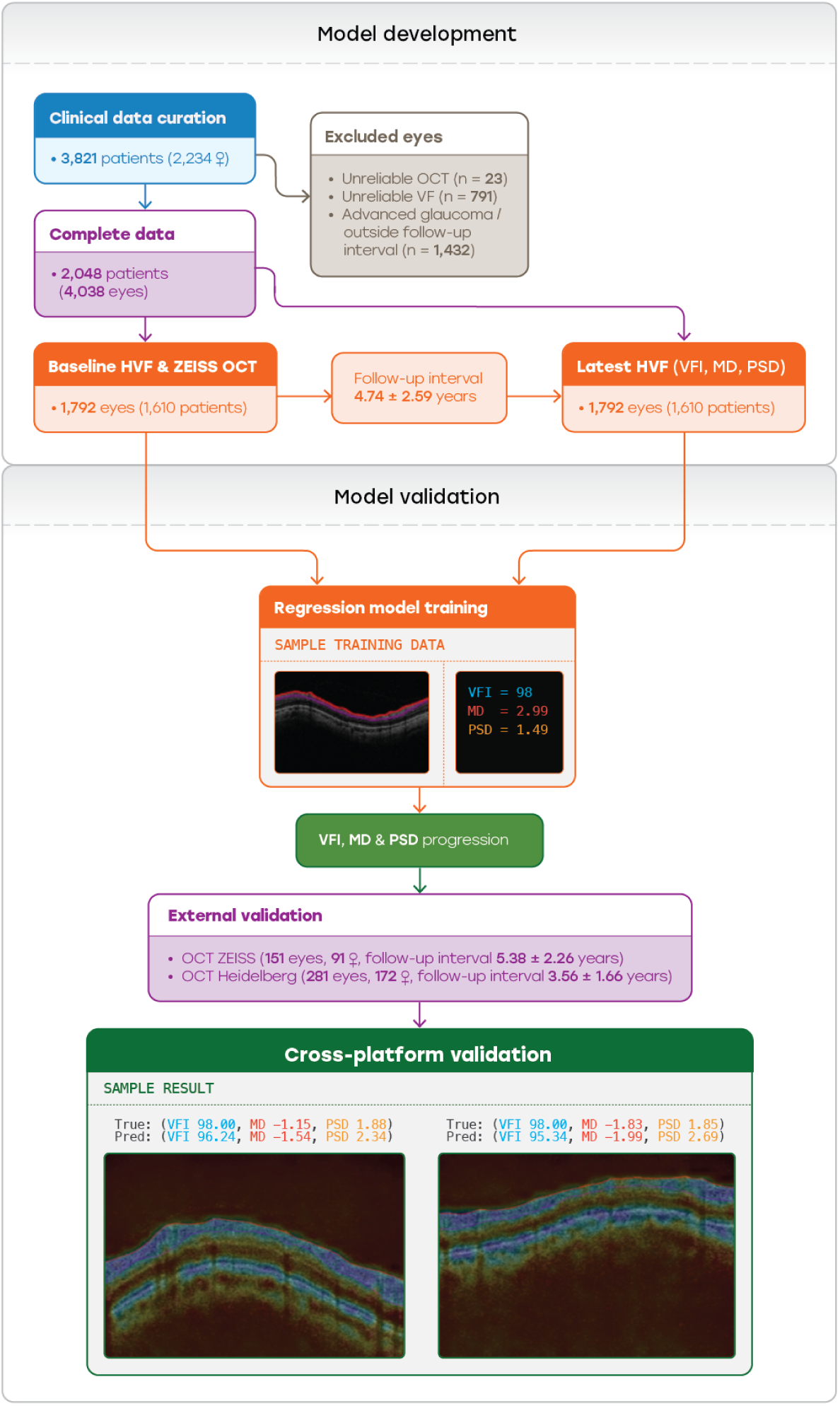
The study outlines the model development and validation phases across datasets from three different clinics.

### Patient Selection

For model training, all participant data were extracted from Hobart Eye Surgeons between May 2009 and September 2019. The glaucoma or suspected patients were classified by an ophthalmologist based on clinical examinations, such as VF and OCT reports. We retrieved data from 4038 eyes (2048 participants) with baseline and latest follow-up VF test reports. Participants with unreliable VF tests, poor-quality images, artefacts of the baseline OCT scans (n=23), or unreliable VF (n=791) were excluded, resulting in a total of 3,224 eyes. We ensured optimal model training for predicting future functional vision. The patient cohort was curated based on follow-up intervals, specifically targeting those with follow-up ranging from 2.15 to 7.33 years (mean ± standard deviation: 4.74 ± 2.59 years). This careful selection was essential to minimise the influence of outliers arising from a limited dataset, ensuring our training process was both robust and reliable. Besides, we included only participants who had both baseline VF and OCT-RNFL scans of the ONH, and finalised follow-up data for model development.

Essendon and Gladstone Park datasets were selected for external validation to assess the model’s performance across different OCT systems (Carl Zeiss and Heidelberg Spectralis). The Essendon dataset consisted of images acquired using the Carl Zeiss OCT system and was used as external validation. A total of 151 reliable OCT-RNFL scans (out of 188 scans) from 92 patients were extracted on the same day of the baseline VF testing or within a time frame of ± 6 months. The Gladstone Park dataset consists of images acquired using the Heidelberg Spectralis OCT system and was also utilised as an external validation set to evaluate the robustness and generalisability of the model when applied to data from a different OCT device. Out of 377 scans, a total of 281 OCT-RNFL scans from 166 patients were selected.

### Humphrey Visual Field Testing

All participants underwent VF testing using SAP with a 24-2 Swedish Interactive Threshold Algorithm (SITA) fast or standard protocol (Humphrey Field Analyzer 2 or 3, Carl Zeiss Meditec, Inc.). We extracted various parameters from the baseline and the follow-up VF reports. These parameters included the patient’s age, sex, and the date of OCT, false negatives (FN), false positives (FP), fixation losses (FL), VFI, MD, PSD, and glaucoma hemifield test (GHT). Additionally, we recorded the time intervals between the baseline and follow-up in years. The reliability criteria for acceptable VF reports included having FL, FP, and FN of 33% or lower.^26^

### Spectral-Domain OCT-RNFL Scanning

Peripapillary RNFL imaging was performed using the Cirrus 6000 spectral-domain OCT (SD-OCT) scanner from Carl Zeiss Meditec, Inc. at Hobart Eye Surgeons and Essendon Eye Clinic. The device acquires high-resolution volumetric data using a 6 x 6 mm optic disc cube scan, consisting of 200 A-scans per B-scan and 200 B-scans. The OCT-RNFL scans from patients were extracted on the same day as the baseline VF testing or within ±6 months. The OCT scans were reviewed directly on the ZEISS FORUM platform, which integrates OCT data with other imaging modalities for synchronised longitudinal assessment.^19^ Visualisation can be shown in full colour or grayscale, with optional overlays including segmentation boundaries, RNFL deviation maps, TSNIT (Temporal, Superior, Nasal, Inferior, Temporal) plots, and en-face fundus-aligned projections. The OCT-RNFL scan was extracted from the ONH region using the software’s built-in features with fixed brightness and contrast values of 255 and 30, respectively, to maintain consistency. The scans obtained were saved in bitmap (BMP) format with a resolution of 1356 x 904 pixels. Images with a signal strength of 7 or higher were included, while scans with artefacts, segmentation errors, or indications of patient movement were excluded.

The Heidelberg Spectralis SD-OCT was used to acquire OCT-RNFL circular B-scans at Gladstone Park Eye Clinic. The scan protocol used was a circular line scan centred on the ONH, with a 3.5 mm diameter circle. The OCT-RNFL scan was extracted from the ONH region using the Heidelberg Eye Explorer (HEYEX)^27^ software’s built-in features with fixed brightness and contrast values of 255 and 30 (respectively) to maintain consistency. The scans obtained were saved in BMP format with a resolution of 1356 x 904 pixels. Images with a quality score of 20 or higher (out of 40)^28^ were included, while any scans with artefacts, segmentation errors or indications of patient movement were excluded.

### Data Refinement for Forecasting Model

All patients under 18 years of age, patients with ocular or systemic conditions known to affect visual fields, such as retinal detachment and optic neuritis, and those with missing VF indices or OCT data were also excluded to ensure the completeness of the dataset and prevent errors in model training and evaluation.

We further filtered out advanced glaucoma, excluding eyes with VFI, MD, or PSD worse than 79%, −12 dB, and 8 dB, respectively. The MD cut-off is chosen to reflect the Hodapp-Parrish-Anderson mean deviation classification.^29^ This exclusion focused the model on early to moderate glaucoma, where subtle structural changes (e.g. RNFL thinning) are more challenging to detect and predict. Advanced glaucoma cases often exhibit a flooring effect in RNFL measurements, where further thinning is undetectable due to involvement of non-neural components (e.g. glial tissue).^30^

In addition, to ensure consistency in longitudinal modelling, we applied a filtering step to patient follow-up intervals. As shown in **Figure 2**, the distribution of follow-up intervals was examined, and data were restricted to those within a standard deviation of the mean (4.74 ± 2.59 years), corresponding to a range of 2.15-7.33 years. This range was chosen to reflect typical clinical follow-up patterns for glaucoma patients, who are recommended to undergo VF and OCT assessments every 3 to 6 months.^31^. As a result, those with follow-up intervals outside of the recommended time frame were excluded, and 1,792 eyes were captured, representing the central majority of patients.

**Figure 2:**
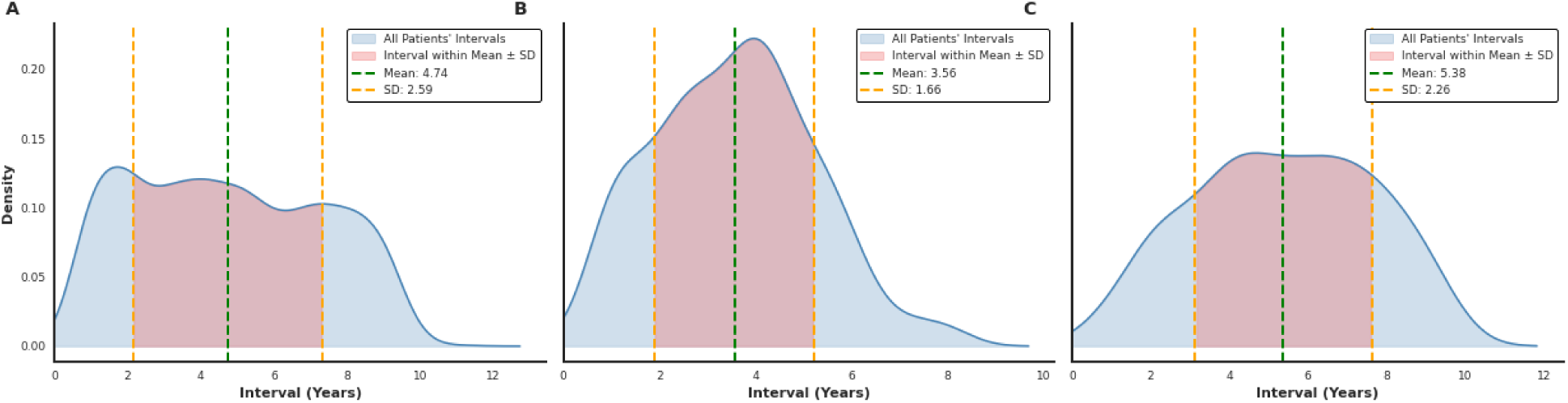
Distribution of follow-up intervals across three OCT datasets were used for model development and external validation. Kernel density estimation (KDE) curves show the distribution of visual field follow-up intervals between baseline and follow-up across the datasets. The blue curve represents all intervals, with the red curve highlighting those within the mean ± standard deviation. A) Model development (OCT ZIESS), B) external validation cohort (OCT ZIESS), C) external validation (OCT Heidelberg).

### Deep Learning Architecture and Training

To identify the optimal model configuration, we utilised the fastai framework^32^ to train multiple variations of possible deep learning architectures (n=135). This included a combination of 15 basic deep learning models, three different loss functions, and three different optimisers (Stochastic Gradient Descent (SGD), Adam, and Ranger)^33^, while keeping other parameters constant to isolate the best-performing combinations of the three (**Supplementary Table S1)**. Each model was trained for limited epochs using varying batch sizes (16, 32 and 64). Batch sizes were selected to balance computational efficiency with model stability, and training for each configuration was limited to five epochs during the architecture screening phase to allow adequate learning while minimising the risk of overfitting.

We extensively implemented data augmentation and normalisation techniques (**Supplementary Table S2**) before training these models to broaden and diversify our training dataset. The dataset was randomly split, with 80% for training and 20% for validation. The chosen loss function for training the models was MSELossFlat(), which calculates the Mean Squared Error. During the training and validation phases, we monitored two regression metrics, Root Mean Squared Error (RMSE) and Mean Absolute Error (MAE), to assess the models’ performance, offering insights into the model’s prediction errors. The original BMP images (1356 x 904 pixels) were downsized to 518 x 518 pixels as input into the model.^34^

All models underwent a two-step training process to attain optimal results. Initially, these models’ layers were frozen to preserve the pre-trained weights^35^, and the last layer was fine-tuned to our OCT-RNFL dataset for a maximum of 10 epochs at a learning rate of 2e-3, enabling the model to adapt generalised features from the initial task while predicting the VF metrics. The learning rate finder was implemented, which suggested the optimal rates. After this, the entire model was unfrozen, allowing all layers to be trainable. This additional training facilitated more precise adjustments throughout the model to better fit our dataset. Subsequently, our model underwent 10 epochs of training employing the 1-cycle policy.^36^ This approach dynamically adjusted the learning rate and momentum on a per-batch basis, fluctuating rates between 1e-7 and 5e-4. In order to improve the stability of the model and prevent overfitting, we implemented an early stopping feature that would stop the training process if the validation loss did not improve by at least 0.01 for three consecutive epochs. We also applied weight decay (2e-3), a regularisation technique, to prevent overfitting and enhance generalisation.

The model’s explainability was visualised using salience map overlays from the final layer of our ViT model, highlighting which regions of the OCT-RNFL scans most strongly influenced the model’s predictions. ViT processes each OCT scan as a sequence of small patches (14 x 14), and the final attention weights indicate which patches contribute most to the model’s output. These attention values were converted into heatmaps, upsampled to match the OCT resolution, and merged with the original image to construct visual overlays.

### Evaluation Metrics

We used a range of evaluation metrics to assess the performance of our regression model against the actual values in forecasting visual function from a baseline OCT-RNFL scan. The primary metrics used were MAE and RMSE, which quantified the average absolute difference between actual and predicted VF indices, providing insight into the model’s precision. Furthermore, a bootstrap resampling technique was implemented to estimate the 95% confidence intervals (CI) for all these regression metrics.^37^

The experiments were conducted on a virtual Ubuntu desktop with an NVIDIA A100 GPU (40GB of GPU RAM). The Python programming language was executed with various libraries such as PyTorch, Fastai, TorchVision, Matplotlib, and Scikit-learn.^38–40^

## RESULTS

### Dataset Characteristics

The model was primarily developed on the Hobart Eye Surgeons dataset from 1792 eyes from 1610 participants. The mean age at baseline was 68.81 years, and 56.7% of the cohort were female **(Table 1)**. The correlation matrix of VF indices and clinical covariates in the final training dataset is visualised in **Supplementary Figure S1**. The external validation was performed on two devices: Carl Zeiss and Heidelberg Spectralis OCT. **Table 2** highlights the significant (*p*<0.001) differences in global key VF indices between the baseline and follow-up assessments, as determined by the Wilcoxon signed-rank test.^41^ These results indicated that glaucoma had progressed over time. The distribution of VF indices of the training data is visualised through a violin plot (**Supplementary Figure S2).**

**Table 1:** Demographic details from three clinical datasets.

| Cohort | Number of Patients,<br>n (Female %) | Number of<br>OCT Scans | Baseline Age,<br>Mean (SD) in<br>Years |
| --- | --- | --- | --- |
| Model Development | 1610 (56.70) | 1792 | 68.81 (12.51) |
| External Validation<br>(Zeiss OCT) | 92 (60.67) | 151 | 69.93 (13.64) |
| External Validation<br>(Heidelberg OCT) | 166 (59.28) | 281 | 77.08 (9.78) |
| Abbreviations: OCT: Optical Coherence Tomography, SD: Standard Deviation, Zeiss OCT: Zeiss Optical Coherence Tomography (manufacturer), Heidelberg OCT: Heidelberg Optical Coherence Tomography (manufacturer) |  |  |  |

**Table 2:** Comparative analysis of baseline and follow-up VF Indices.

| Visual Field Index | Clinical Dataset | Baseline | Follow-up | P-value* |
| --- | --- | --- | --- | --- |
| **VFI (%)** | Model Development | 91.99 ± 15.06 | 88.14 ± 19.67 | <0.001 |
|  | External Validation (Zeiss OCT) | 96.21 ± 5.56 | 96.27 ± 4.26 | <0.001 |
|  | External Validation (Heidelberg OCT) | 95.06 ± 5.85 | 95.45 ± 4.30 | <0.001 |
| **MD (dB)** | Model Development | -3.02 ± 5.20 | -4.39 ± 6.43 | <0.001 |
|  | External Validation (Zeiss OCT) | -1.56 ± 2.80 | -1.61 ± 2.43 | <0.001 |
|  | External Validation (Heidelberg OCT) | -2.41 ± 3.06 | -2.16 ± 2.32 | <0.001 |
| **PSD (dB)** | Model Development | 3.23 ± 2.93 | 3.81 ± 3.26 | <0.001 |
|  | External Validation (Zeiss OCT) | 2.84 ± 2.32 | 2.53 ± 1.53 | <0.001 |
|  | External Validation (Heidelberg OCT) | 2.88 ± 2.09 | 2.61 ± 1.45 | <0.001 |

**Abbreviations:** **OCT:** Optic Coherence Tomography, **SD:** Standard Deviation, **VFI:** Visual Field Index, **MD:** Mean Deviation, **PSD:** Pattern Standard Deviation, **dB:** decibel, \*The comparison between baseline and follow-up VF indices was made using the Wilcoxon signed-rank test.

### Model Configuration

Our analysis of optimal model configurations revealed that ViT with a self-supervised ViT model using DINOv2^9^ consistently achieved the lowest MAEs, ranging between 5.4 and 7.6. In comparison, CNNs-based architectures, such as DenseNets, had MAEs ranging from 9.0 to 9.6, while ResNet and VGG baselines showed MAEs from 9.8 to 10.3, as indicated in **Supplementary Figure S3**. The best-performing configuration, ViT Small Patch14 using DINOv2 with a batch size of 16, reduced the error by approximately 47% compared to the least effective model among the top ten. These findings highlight the superiority of ViT-based architectures for predicting visual function from a baseline OCT-RNFL scan.

### Performance of Forecasting Model

The ViT-based regression model achieved low forecasting errors for VFI, MD, and PSD on internal and external datasets, across Zeiss and Heidelberg OCT acquisitions (**Table 3**). These findings demonstrate the model’s cross-platform generalisability in predicting key VF indices. However, it is important to emphasise that these results focus on predicting VF indices rather than diagnosing glaucoma or specifying the onset of clinically significant vision loss. For VFI, the model attained an MAE of 3.32% on the internal validation set, indicating the average deviation between actual follow-ups and predicted VFI values. When predicting MD, the observed errors were considerably lower, with MAE and RMSE values of 1.78 dB and 2.25 dB, respectively. The model demonstrated its highest precision in estimating PSD, with an MAE of 1.10 and an RMSE of 1.50.

**Table 3:**
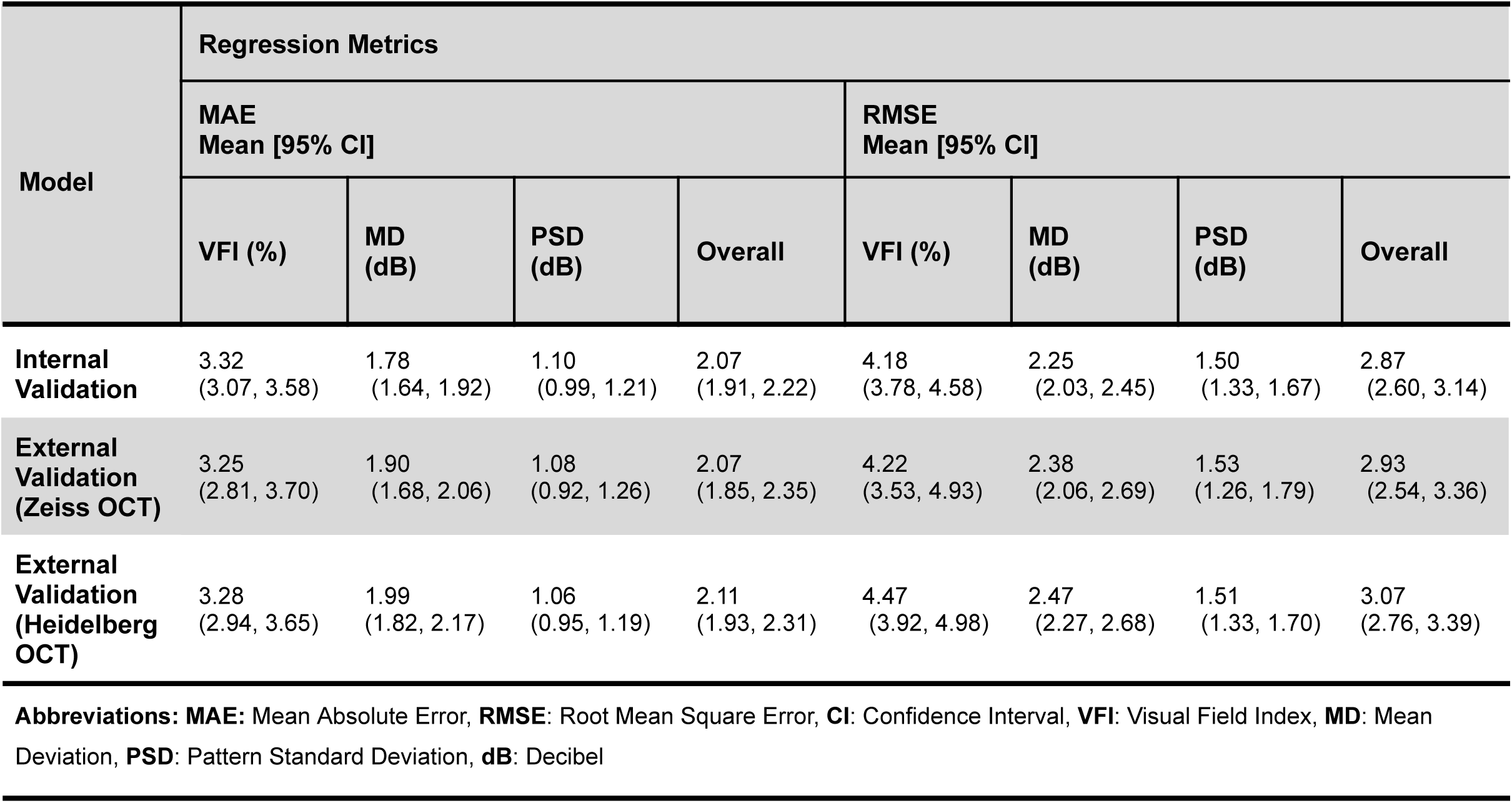
Evaluation of the model’s performance across Zeiss and Heidelberg OCTs.

When the model’s overall performance on combined metrics (VFI, MD, and PSD) was evaluated, the MAE was 2.07, and the RMSE was 2.87. The model achieved comparable accuracy across datasets. Internal validation yielded an MAE of 2.07 (95% CI: 1.91-2.22), while external validation showed 2.11 (95% CI: 1.93-2.31) for Heidelberg OCT and 2.07 (95% CI: 1.85-2.35) for Zeiss OCT (**Table 3**). Substantial overlap of the bootstrapped confidence intervals indicates no statistically significant difference in model performance between internal and external cohorts.

Bland-Altman plots were generated to visualise the model’s predicted three key VF indices in **Figure 3**, compared with actual and predicted values across both datasets, Zeiss and Heidelberg OCTs. The red dashed line shows the mean bias, and the green dashed lines show the 95% limits of agreement. For Heidelberg OCT, the mean bias (95% limits of agreement) was −0.83 (−9.45 to 7.80) for VFI, 0.02 (−4.83 to 4.88) for MD, and 0.22 (−2.73 to 3.16) for PSD. For Zeiss OCT, the corresponding values were 0.32 (−7.95 to 8.60), 0.14 (−4.54 to 4.81), and 0.14 (−2.85 to 3.13), respectively, indicating reasonable agreement between predicted and observed functional outcomes across devices.

**Figure 3:**
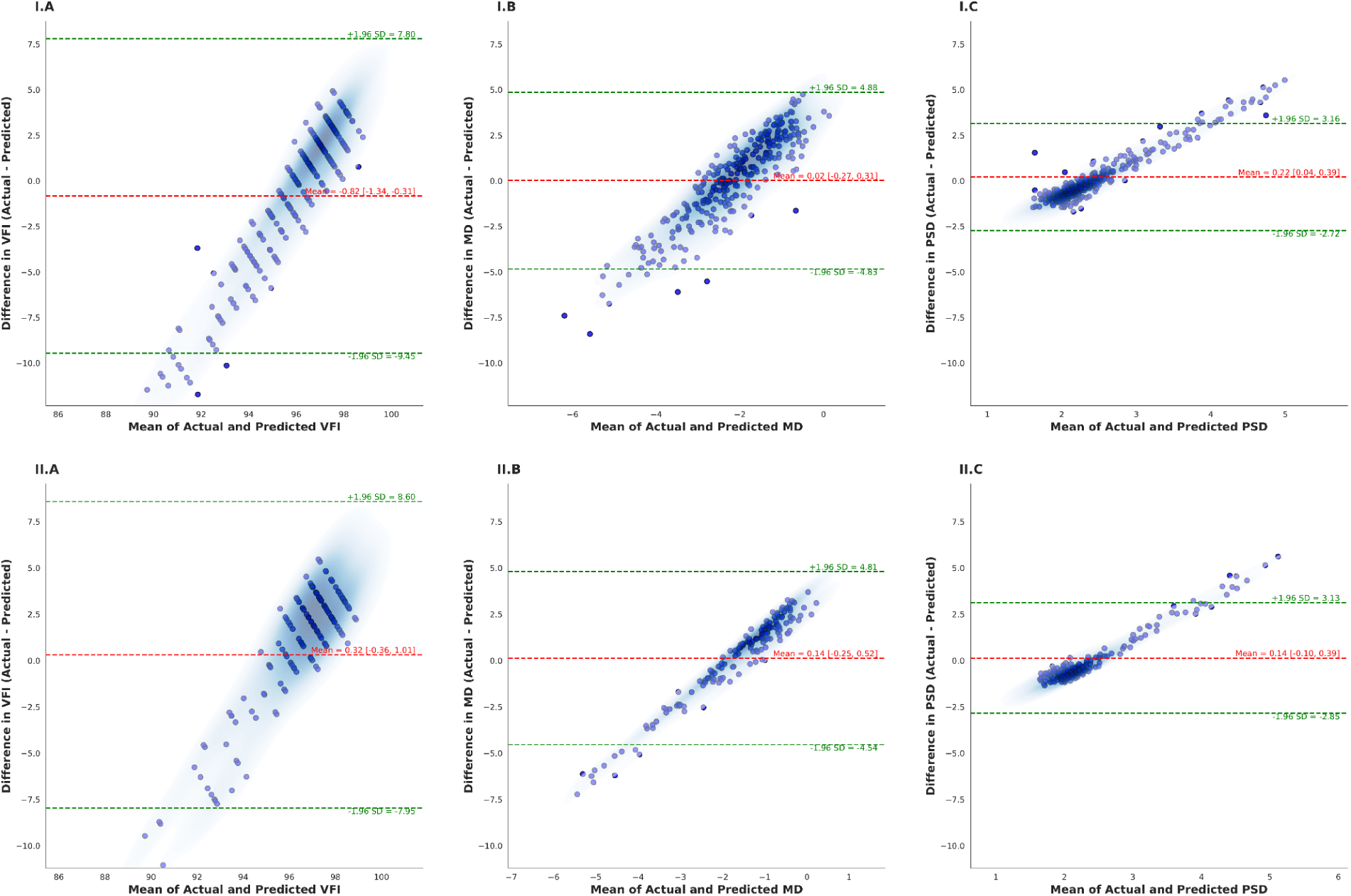
Bland-Altman Plots showing agreement between actual and model-predicted visual field indices for two independent external OCT datasets. Columns A–C show visual field index (VFI), mean deviation (MD) and pattern standard deviation (PSD), respectively. The top row (I.A–I.C) corresponds to the Heidelberg OCT validation dataset and the bottom row (II.A–II.C) to the Zeiss OCT validation dataset. The red dashed line denotes the mean difference between actual and predicted values, and the green dashed lines indicate the 95% limits of agreement (mean ± 1.96 SD).

### Saliency-Based Interpretability

Saliency maps highlighted the model’s decision-making areas in RNFL-OCT scans (**Supplementary Figures S4-S6**), providing interpretability by visualising the anatomical regions most influential in predicting functional indices (VFI, MD and PSD) across internal and external datasets. The highlighted areas in blue consistently localise to the RNFL and inner retinal layers (upper blue band in each image), which are structurally linked to functional vision loss and thus represent clinically meaningful features. In addition, the retinal pigment epithelium (RPE) and outer retinal nerve layers (lower blue band in each image) were also highlighted. The model’s predictions align closely with true values, and the heatmaps correspond well to areas showing structural integrity or subtle thinning of RNFL. This underscores the model’s capability to capture biologically relevant patterns from the OCT-RNFL scans. Clinically, these Saliency maps clearly demonstrate the connection between structural OCT features and predicted overall VF indices, ensuring the model emphasises relevant glaucomatous regions of RNFL OCT scans.

## DISCUSSION

We developed and validated a ViT-based regression model to forecast the functional impact of glaucoma progression (measured by VF indices) from a single circular B-scan of OCT-RNFL at ONH. Our model was shown to predict progression up to seven years before the worsening of glaucoma. To the best of our knowledge, this approach has yet to be reported in the literature. In contrast to previous studies, our research leverages a ViT-based regression model, which is quite novel as the literature is dominated by CNNs-based models.^20,42^

Importantly, the accuracy of our ViT-based predictions falls within the range of inherent clinical variability observed in standard automated perimetry. Swanson et al. reported that when the same individual is tested twice on Humphrey Visual Fields, MD varies between +2.5 and −1.4 dB (mean 0.0, SD 0.9 dB) while PSD ranges from 0.9 to 2.1 dB (mean 1.5, SD 0.3 dB).^43^ Our model achieved MAEs of 1.78 for MD and 1.10 for PSD, which are comparable to or lower than the inherent test-retest variability. This suggests that the model’s predictions were both statistically robust and clinically meaningful, as it captures visual function within the limits of current measurement reproducibility. This precision enables the identification of eyes requiring treatment escalation from the baseline visit, which is critical in clinical settings with high loss to follow-up. The model may therefore support timely intervention to prevent irreversible vision loss from glaucoma.

Our model outperformed previous studies in predicting VF indices from OCT scans, achieving the lowest MAE and RMSE.^22,23,44–48^ For example, Maetschke et al (2019)^22^ estimated the VFI and MD from the OCT volume with an RMSE of 12.0±1.61 and 4.1±0.44, respectively, whereas our model forecasted the VFI and MD values with an RMSE of 4.18 (95% CI: 3.78-4.58) and 2.25 (95% CI: 2.03-2.45), respectively on the model development cohort (internal validation). They observed a marginally higher accuracy when using ONH scans as inputs to the model compared to macula scans^22^, additionally, our study only used the baseline OCT-RNFL scans of ONH as input for forecasting the model. Another artificial neural network (ANN) model, trained on diverse OCT-derived RNFL datasets, acquired an MAE of 4.0 dB (95% CI: 3.8-4.2) and an RMSE of 5.2 dB (95% CI: 5.1-5.4) on the testing dataset. The MAE varied from 3.3 to 5.9 dB and the RMSE from 4.4 to 8.4 dB on independent datasets.^49^ Unlike those models, our model demonstrated superior performance in forecasting VF indices utilising only the raw OCT-RNFL B-scans of ONH. Our top-performing model achieved overall MAE and RMSE of 1.40 and 1.74, respectively, for forecasting visual function (VFI, MD, and PSD). However, direct comparison of the two studies is not possible due to their differing data sets and model architectures.

A ViT-based regression model may outperform CNNs-based approaches for several important reasons related to the nature of OCT-RNFL imaging. First, OCT B-scans contain high-resolution structural information. In our dataset, each scan is 1356 × 904 pixels before downsizing, whereas most CNNs-based architectures are designed for smaller inputs (typically 224 × 224 pixels), especially when pretrained. Downsampling to fit CNNs’ input requirements can result in loss of fine structural details, which are critical for detecting subtle RNFL thinning in early glaucoma. In contrast, ViTs process images as a sequence of patches (in our case, 14 × 14-pixel patches), allowing the model to preserve spatial detail across the entire image and capture long-range relationships between regions of the ONH that CNNs may miss due to their localised convolutional kernels. Additionally, the self-attention mechanism in ViT enables global contextual understanding instead of focusing solely on local features.^50^ This makes it particularly effective at identifying subtle patterns that correlate with functional visual field loss. In addition, our study benefits from a substantially larger sample size than most prior regression studies, providing the ViT model with a more diverse set of training examples. This larger dataset may have improved the model’s ability to generalise across different patient populations and reduce the risk of overfitting. This may have enhanced reliability in predicting VF indices from OCT-RNFL scans.

The ViT model was externally validated on two smaller datasets using Zeiss OCT and Heidelberg OCT, with performance comparable to that in internal validation. External validation across both Zeiss and Heidelberg OCT datasets demonstrated error rates highly consistent with those observed in the model development cohort, indicating stable performance across devices and populations. Importantly, the overlap in bootstrapped confidence intervals indicates no statistically significant difference in model performance across datasets, suggesting the model’s robust generalisability despite differences in OCT devices. This finding was surprising and contrasts with existing literature, which agrees that models trained on images from one OCT machine may lead to instrument-specific biases that reduce performance in a different clinical setting.^51^ Our model’s consistent performance across Carl Zeiss and Heidelberg Spectralis devices likely stems from standardised preprocessing (e.g. standardised brightness/contrast), which may have reduced heterogeneity. Another possible reason is that the ViT model’s attention mechanism captured the most relevant anatomical areas for training and therefore produced more robust results than CNNs-based models. Another factor may be that regression models are inherently more generalisable than classification models, since they predict continuous VF indices (MD, VFI, PSD) rather than discretising progression into classes (stable vs progressing). This may avoid information loss and allow the model to learn subtle trends across the entire spectrum of disease severity. On the other hand, classification tasks often require setting cutoffs (e.g., defining MD > −6 dB as “mild” glaucoma), which vary across studies and populations. These strict, sometimes arbitrary thresholds may reduce the model’s generalisability and limit its clinical utility.

In terms of limitations, our study’s focus on using OCT-RNFL scans to predict VF indices may oversimplify the multifaceted nature of POAG progression, which is influenced by a complex interplay of factors beyond OCT structural features. Clinical variables such as age, sex, baseline intraocular pressure, central corneal thickness, and systemic conditions such as diabetes or hypertension are risk factors for POAG, but we did not include them as model inputs in this study. Similarly, genetic factors and lifestyle variables such as smoking^52^ or medication adherence^53^ were not considered, as well as the effectiveness of treatment, which may further limit the model’s ability to reflect real-world complex interactions. However, the factors mentioned above can be easily obtained from the patient during the baseline first visit and could therefore be integrated into the model to improve performance. Numerous studies demonstrate that a multimodal input approach increases predictive accuracy for classifying POAG progression.^54–56^ Our main utilisation of this model is in glaucoma clinics; thus, the presence of progression does not imply treatment failure, but rather highlights the heterogeneous nature of glaucoma and the need for personalised risk assessment to identify eyes that may require closer monitoring or more intensive intervention.

In conclusion, our ViT-based regression model has the potential to predict key VF indices (VFI, PSD and MD) from a single baseline OCT-RNFL scan before the onset of glaucoma. The model outperforms existing CNNs-based models and provides interpretable saliency maps highlighting RNFL and inner retinal regions, consistent with glaucoma pathophysiology. This offers a tool with potential clinical utility for identifying patients with glaucoma likely to progress more rapidly, enabling timely, personalised management. Our model could be integrated into existing OCT systems to assist clinicians to better patient outcomes. This study also provides a reliable, objective method for predicting VF indices, which are essential for monitoring glaucoma progression and tailoring treatment without requiring long-term follow-up.

## Data Availability

All data produced in the present study are available upon reasonable request to the authors

## Acknowledgements

This work was supported by an Australian National Health and Medical Research Council Leadership Award (A.W.H.). Grammarly was used to improve readability and grammar.

