## Supplemental information for "Predicting visual function before glaucoma onset from baseline optical coherence tomography scans using deep learning"

**Supplementary Table S1:** *The combination of model architectures and parameters was selected during screening.*

| Model Architectures | Optimisers | Loss Functions |
|---|---|---|
| • Vit_small_patch14_dinov2<br>• Vit_small_patch14_reg4_dinov2<br>• Vit_base_patch14_dinov2<br>• Vit_base_patch14_reg4_dinov2<br>• Resnet18<br>• Resnet34<br>• Resnet50<br>• Resnet101<br>• Resnet152<br>• Densenet121<br>• Densenet169<br>• Densenet20<br>• Densenet161<br>• Vgg16_bn<br>• Vgg19_bn | • Adam<br>• SGD<br>• Ranger | • MSELossFlat()<br>• L1LossFlat()<br>• nn.SmoothL1Loss() |

**Supplementary Table S2:** *Data transformation and augmentation techniques were used during the development.*

| Technique | Specification |
|---|---|
| Resize | Size = 518 |
| Random Scale | min_scale=0.8 |
| Lightning transform | max_lighting=0.05 |
| Horizontal Flip, Vertical Flip | do_flip=True, flip_vert=False |
| Rotation | max_rotate=20 |
| Warp | max_warp=0.05 |
| Affine Transforms | p_affine=0.75 |
| Zoom | max_zoom=1.05 |

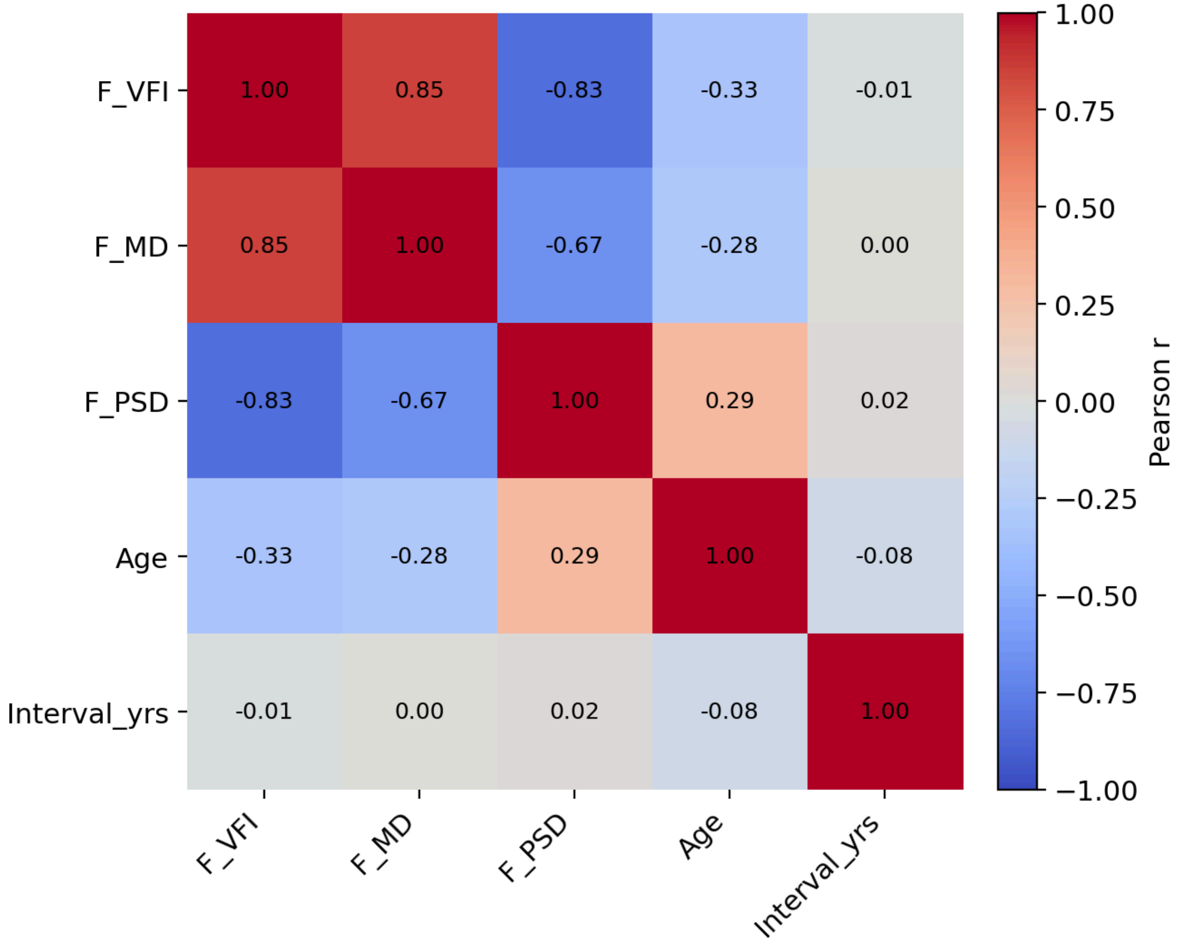


**Supplementary Figure S1:** *Heatmap showing pairwise Pearson correlation coefficients (r) between the latest follow-up visual field index (F_VFI), mean deviation (F_MD), pattern standard deviation (F_PSD), patient age, and the interval between baseline visual field and latest follow-up of visual field testing in years (Interval_yrs).*

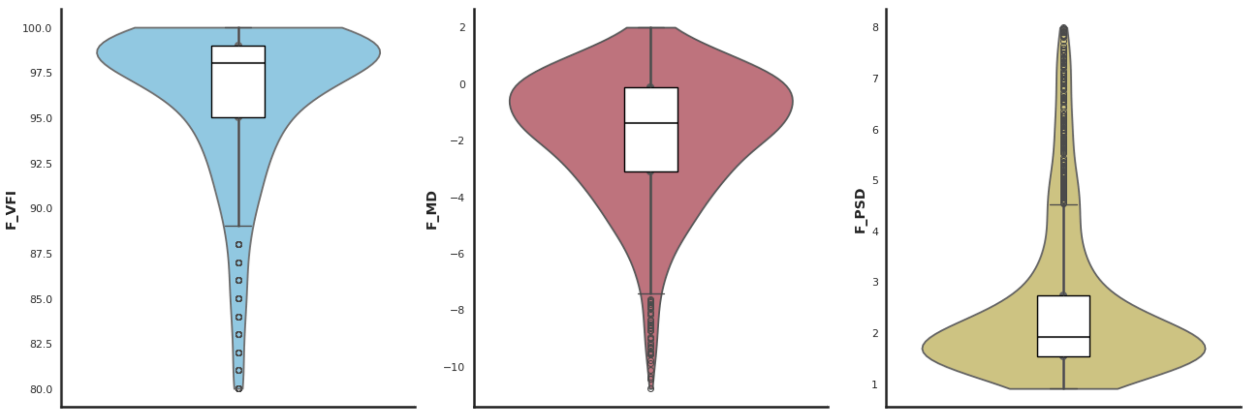


**Supplementary Figure S2:** *The distribution of the final dataset used for model development utilised three global visual field indices (VFI, MD, and PSD) across a mean follow-up period of 4.74 years for analysis.*

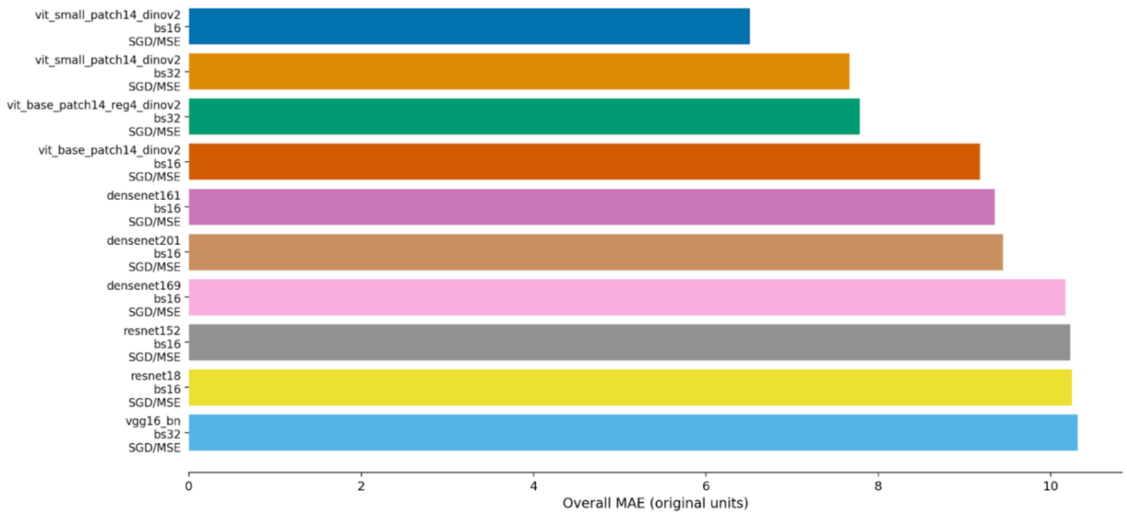


**Supplementary Figure S3:** *Performance of the top ten models was visualised across different combinations of model name, optimiser, and batch size, with the lowest MAE after training for five epochs.*

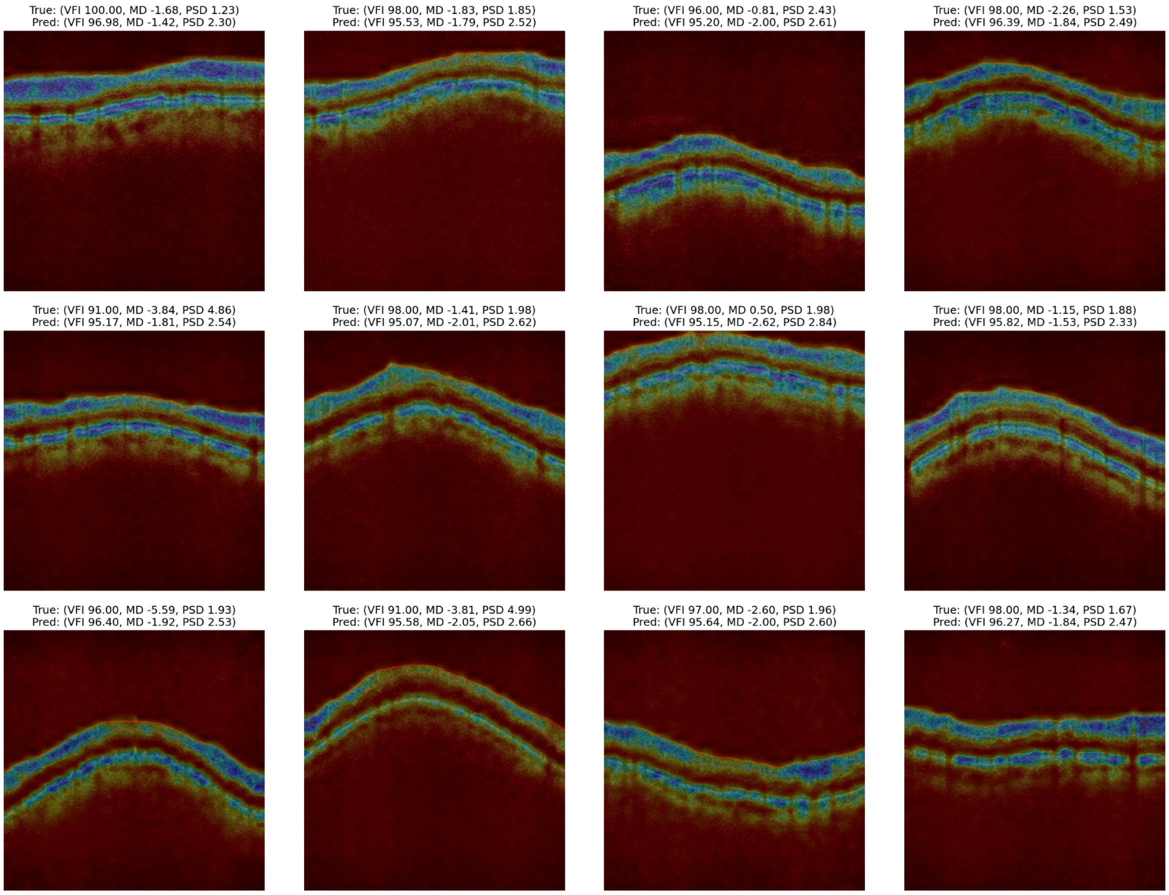


**Supplementary Figure S4:** *Model-derived Saliency maps overlaid on optical coherence tomography (OCT) scans for the internal validation set. Areas highlighted in dark blue denote higher model attention, while green indicates lesser contribution, and black indicates minimal contribution. For each scan, ground-truth clinical metrics are shown above (True) and model-predicted values are reported below (Pred).*

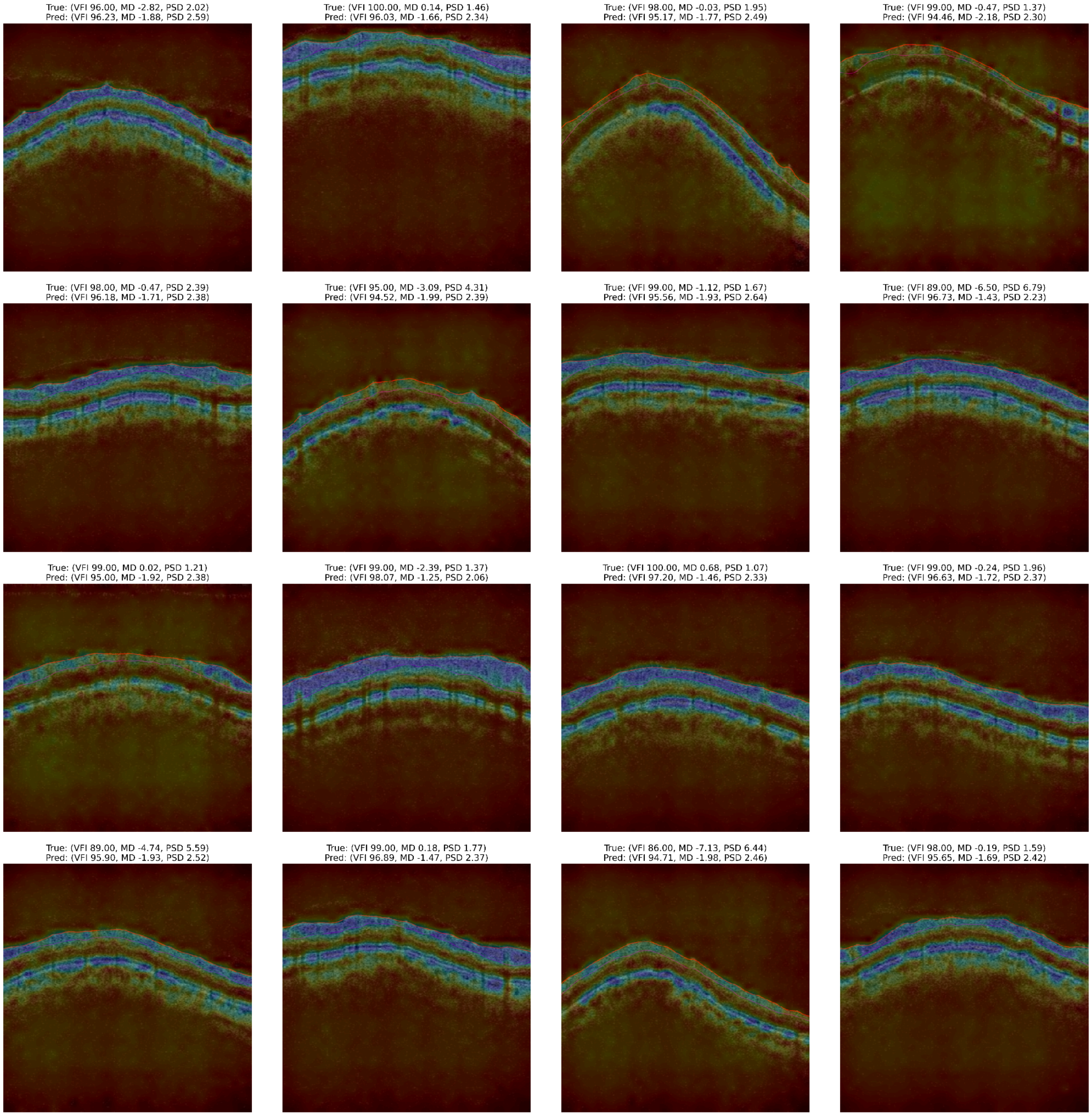


**Supplementary Figure S5:** *Saliency maps derived from the model are overlaid on optical coherence tomography (OCT) scans for the external validation set from the Essendon Eye Clinic (*Carl Zeiss*).*

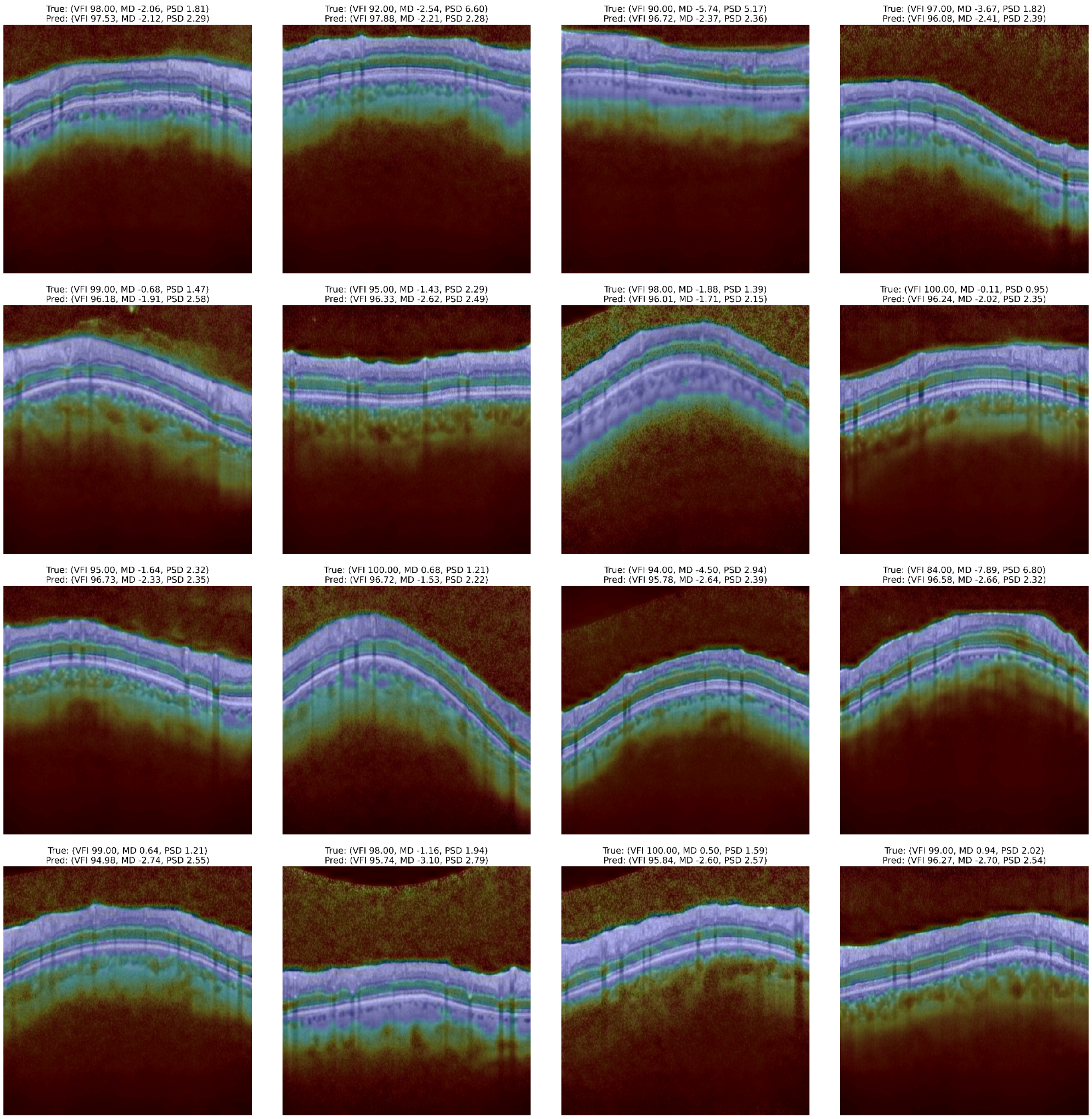


**Supplementary Figure S6:** *Model-generated saliency maps overlay on optical coherence tomography (OCT) scans for the external validation cohort at Gladstone Park Eye Clinic (*Heidelberg Spectralis*).*
